# Demography, hygiene and previous disease prevalence as plausible risk factors associated with Covid-19 deaths across Indian states

**DOI:** 10.1101/2023.06.22.23291740

**Authors:** Bithika Chatterjee, Shekhar C. Mande

**Author notes:** Corresponding author: ^b^Shekhar C. Mande Bioinformatics Centre, Savitribai Phule Pune University, Pune and National Centre for Cell Science, NCCS Complex, Ganeshkhind, Pune- 411007.

## Abstract

Severity of Covid-19 diseases has been disproportionate with higher case-fatality ratio affecting developed nations. In India, states with higher income have reported more number of deaths compared to lower income states. The global burden of diseases India 2019 and the National Health Profile 2019 data was used to draw correlations with Covid-19 mortality at two different dates of peak Covid-19 cases in India. We explored correlation of mortality in different states of India with prevalence of different diseases, demography, development, sanitation etc. The study found a positive correlation with known demographic parameters such as percentage of elderly population(spearman correlation coefficient(rho) =0.44 and 0.46 with 1^st^ and 2^nd^ peak respectively). Similarly, percentage urbanization was seen to correlate well with mortality(rho=0.71 and 0.57) suggesting Covid-19 to be a predominantly urban disease. Prevalence of Autoimmune diseases, and Cancer show higher correlation with deaths. A surprising positive correlation emerged between improved sanitation parameters, such as closed drainage and indoor toilets, with COVID-19 deaths. Overall the multivariate regression model achieved by combining demography, sanitation, autoimmune diseases and cancer gave us the best prediction for Covid-19 mortality(adjusted R square value of 0.71 with peak 1 and 0.85 with peak 2). Analysis of the Covid-19 related data seems to indicate that as the wealth of a state increases, the state’s urban landscape changes often leading to better sanitation facilities. The lifestyle and prevalence to autoimmune diseases as well as cancer also increases. However, this may affect the state’s ability to fight pandemics due to lower exposure to pathogens and immune training.

## Introduction

COVID-19 pandemic has affected millions of people around the world and has caused a significant number of deaths[1]. The rapid spread of the virus and its high transmission rate have put immense pressure on healthcare systems worldwide, leading to a devastating loss of life and economic disruptions. While the virus affects individuals of all ages, certain groups such as the elderly[2,3], those with pre-existing conditions, and frontline workers have been known to be at a higher risk of severe illness and death[4]. Understanding the factors that contribute to COVID-19 outcomes is crucial in formulating effective public health policies and interventions to mitigate the impact of the pandemic.

In our previous study we had attempted to understand the factors that lead to varying mortality observed in different countries and we had noticed a specific pattern emerging globally. Countries with better access to sanitation and with improved demographic parameters such as life expectancy, higher GDP and higher burden of autoimmune diseases appeared to cause more Covid mortality[5]. It is already a known fact that the virus affects individuals differently with some comorbidities and underlying conditions having been identified as risk factors for severe illness and death[6]. Indeed many studies have highlighted lifestyle related conditions such as obesity, high blood pressure, diabetes and even cancers to be higher risk factors for Covid mortality[7–9]. We attempted to analyze prevalence of health and sanitary conditions in different states of India, and attempted to observe if any correlations emerge with respect to Covid-19 mortality.

Our analysis of data from developed countries and Indian states shows a surprising correlation between certain sanitation parameters and COVID-19 deaths, particularly those related to closed drainage and indoor toilets. We propose that this might be explained by evidence indicating that COVID-19 is airborne[10] and can spread inside households from toilets, even while patients are in isolation[11–13]. Additionally, we found that exposure to previous infections with viruses, bacteria may confer a protective immune response against COVID-19, whereas autoimmune diseases and certain lifestyle diseases could aggravate an already elevated inflammatory response in the body, potentially increasing the risk of severe illness and death. This observation goes in line with the hygiene hypothesis that postulates that the more pathogens that a person encounters in one’s lifetime the more robust their immune system becomes[14,15]. This could partly explain lower fatalities in African and Asian countries where the population has been exposed to many tropical diseases and parasitic infections[16]. Our findings have important implications for public health policies and highlight the need for further research on the relationship between sanitation, immune response, and COVID-19 outcomes.

### Methodology

The state wise Covid-19 mortality was collected from https://www.covid19india.org. It was collected at approximate dates of peak covid cases in India as indicated from World Health Organization[17]. Although the cases surged at three time points, we proceeded to analyse deaths only for the first and the second peak as by the time the third peak came, most of the population in India was vaccinated[17]. The first peak occurred around 17^th^ September 2020 and second one on 7th May 2021. We normalized the cumulative deaths occurring on both the days by the state and Union Territory’s (UT) total population[18] (Census 2011) to get deaths per million (DPM). The parameter, Covid-19 deaths per million, was chosen instead of cases or case-fatality ratio since it is a more reliable parameter. We removed Telangana as no census data is available for this state in 2011. Moreover, many of the parameters were missing for this newly formed states. Mizoram state was also removed since it did not report a single death at one of the peaks of Covid-19 cases. We collected and analysed data for a total of 26 Indian states and 2 Union territories.

The parameters population density, gender ratio, urban population percentage was obtained from the 2011 Census of India[19]. The percentage of literacy in each state and UT was obtained from the Office of the Registrar General and Census Commissioner[20]. The GSDP(2018-19)/capita was obtained from https://statisticstimes.com/economy/india/indian-states-gdp-per-capita.php. Good Governance Index for year 2019 was collected from http://data-analytics.github.io/Good_Governance/. The prevalence of communicable and non-communicable diseases was taken from global burden of diseases India 2019 https://vizhub.healthdata.org/gbd-compare/india#0. This prevalence percentage of the disease in each state was standardized by age and obtained for both the genders. The percentages of infant vaccination parameters, slum household, below poverty line population, sanitation and expenditure on health hospital beds was obtained from National Health Profile 2019 14^th^ issue https://www.cbhidghs.nic.in/. Since some parameters were missing for the newly formed state Chhattisgarh we imputed the value of Bihar as the states have comparable variables.

### Statistical Analysis

We calculated (Spearman) correlation coefficients of different variables with the DPM on 17^th^ September 2020 (DPM1) and 7^th^ May 2021 (DPM2) to obtain certain selected parameters that might be associated with Covid-19 mortality. We used a cut-off of 0.40 for positive and negative correlation coefficients to select our parameters for multivariate linear regression analysis. We then combined similar variables to obtain adjusted R square values with DPM to depict how much percentage of variability can be explained by each of the broader category variables. We further proceeded to combine the broader variables that show more than 0.40 adjusted R square values for both DPM1 and DPM2. We used statistical tool R for all the statistical analysis.

## Results

WHO has reported 4,46,91,956 confirmed cases of COVID-19 with 5,30,789 deaths as on 16^th^ March 2023 from the onset of pandemic in India[17]. In these three years of pandemic in India, there were three surges of covid cases. The first peak surged in the middle of September 2020 with 6,46,263 cases and 8166 deaths. The second peak surged in early May 2021 with 27,38,957 cases and 28,982 deaths[17]. The first peak was dominated by the milder Alpha variant while the second wave was dominated by the Delta variant which caused more casualty[21].The third peak occurred in Jan 2022 resulting in 7888 deaths due to the milder omicron variant, but good vaccination coverage across the states[17] was achieved by then.

In our analysis we observe that the highest Covid-19 mortality occurred for higher income states like Delhi followed by Maharashtra (Figure 1. (*a*)) whereas states like Bihar and Jharkhand reported relatively lower deaths. In Figure 1(*b*) we see a linear relationship between DPM2 and GSDP per capita of all states with a spearman correlation coefficient of 0.66 (Table1). Such a distinct pattern of covid mortality among the Indian states made us to explore other parameters that could help explain this variation occurring across the states. We therefore analysed many different parameters with respect to deaths per million, including the known risk factors. Results of these are described below:

**Figure 1.**
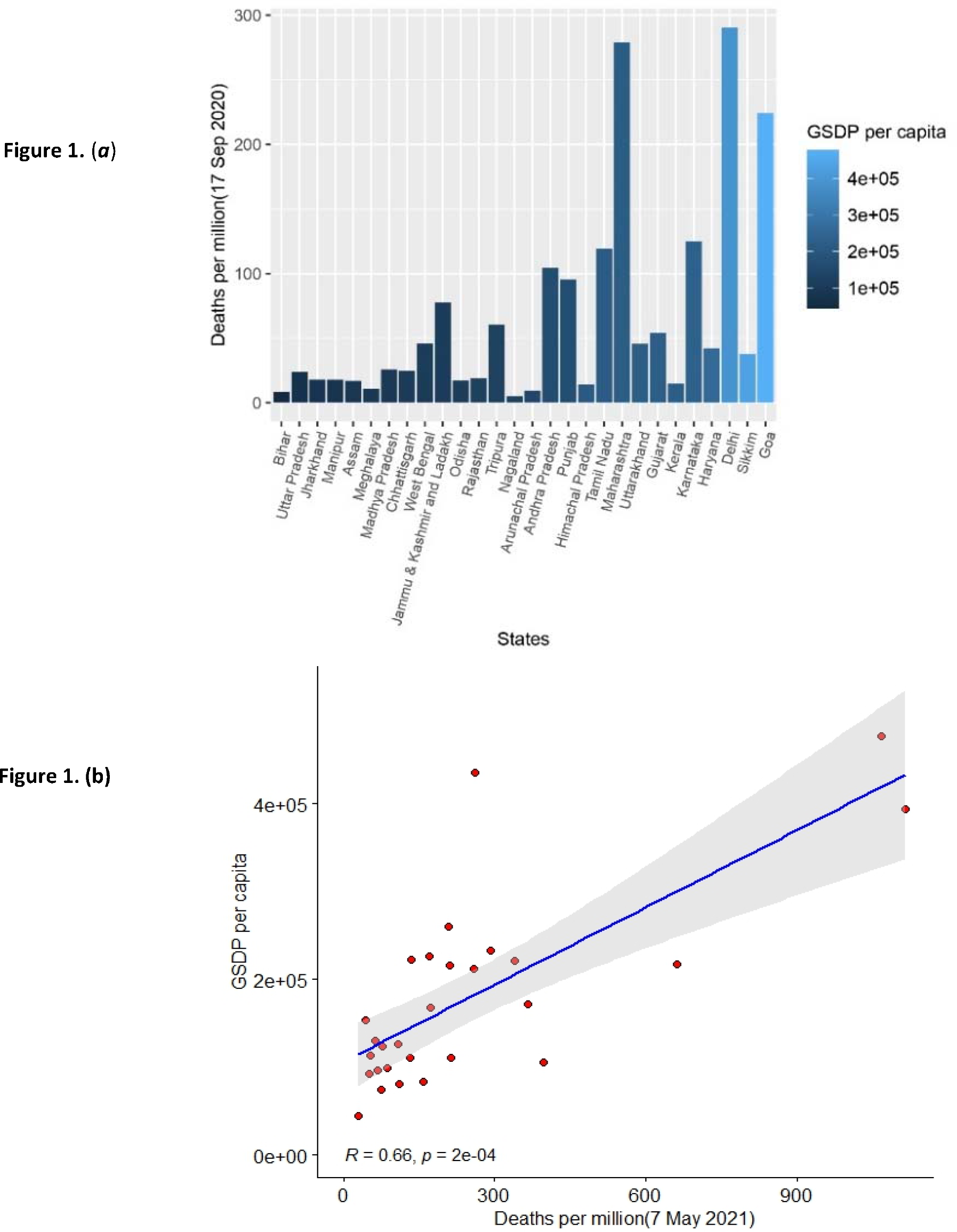
(***a***) Number of deaths per million inhabitants in each state due to COVID-19 as on 17 September 2020. Colour gradient indicates the state’s GSDP per capita for year 2018 to 2019. (***b***) Plot showing positive correlation between Deaths due to COVID-19 as on 7 May 2021 vs. the state’s GSDP per capita for year 2018 to 2019. R denotes Spearman’s correlation coefficient of 0.66. Each dot denotes an Indian state(N=28).

### Demography

The age of elderly population percentage over 60 correlated positively for DPM1 and DPM2 as did the literacy percentage and the Good Governance index. The best correlation was seen with Urbanisation percentage with rho = 0.71 for DPM1 and 0.57 with DPM2 (Table1). Population below poverty percentage was the only parameter that correlated negatively with DPM1 and DPM2 (Table1). Overall the parameters combined excluding GSDP per capita in linear regression gave us an adjusted R^2^ value of 0.67 and 0.51 with DPM1 and DPM2 respectively (Figure 2).

**Figure 2.**
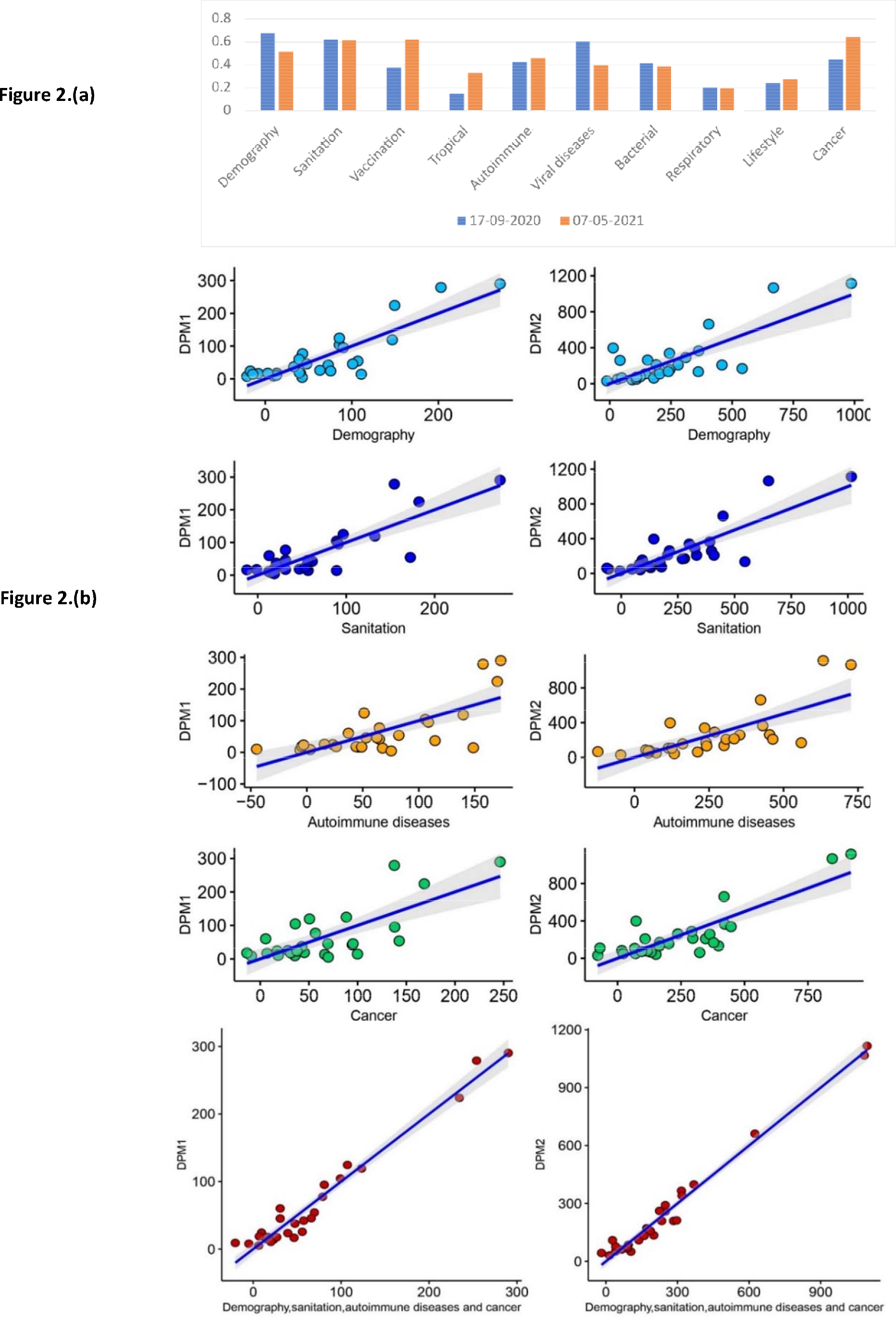
(a) Regression analysis was done with Covid-19 deaths/million in Indian states as on 17-09-2020 and 07-05-2021 as the dependent variable and combinations of different variables; N=28. Graph shows the Adjusted R- squared values depicting how much percentage of variability in the mortality could be explained by the variables in each category. **(b)** Actual values of deaths per million as on 17 September 2020 (DPM1) and 7 May 2021(DPM2) plotted against their predicted values by combining variables of demographics, sanitation, autoimmune diseases and cancer separately and then combining all of them together. Regression analysis was done with DPM1 and DPM2 as the dependent variable and combinations of different variables as explanatory variables; N = 28. Each dot represents an Indian state.

### Vaccination

The universal childhood immunization program in India covers vaccination of BCG, POLIO, DPT, Measles etc. The vaccines coverage percentage of BCG, POLIO, DPT, Measles and Hepatitis correlated positively with the covid deaths while Acute Hepatitis B and Acute Hepatitis E vaccine percentage correlated negatively with DPM1 and DPM2 (Table1). Dosage of Vitamin A during infancy also correlated positively (Table1). Adjusted R^2^ value for combined Vaccination parameters was 0.37 and 0.62 with DPM1 and DPM2 respectively (Figure 2).

### Tropical diseases

Most tropical diseases prevalence in states (Trichuriasis, Leprosy, Hookworm and Ascaris) show negative correlation with DPM except Cystic echinococcus which shows positive correlation (Table1). Adjusted R^2^ value for combined tropical diseases parameters was 0.15 and 0.33 with DPM1 and DPM2 respectively (Figure 2). These correlations as well as regression values were very low and insignificant since most states report very low percentage of these diseases.

### Autoimmune diseases

Prevalence of Gout, Diabetes mellitus type 2 and Inflammatory bowel disease (IBD) correlated positively while Asthma, Psoriasis showed a negative correlation with DPM (Table1). Regression analysis with DPM1 and DPM2 yielded an Adjusted R^2^ of 0.42 and 0.46 respectively (Figure 2).

### Viral diseases

Prevalence of Rabies, Meningitis, Acute hepatitis shows negative correlation with DPM with Measles showing highest negative correlation of -0.57 and -0.73 with DPM1 and DPM2 respectively (Table1). Only prevalence of Dengue showed a positive correlation of 0.6 and 0.4 with DPM1 and DPM2 respectively (Table1). Regression analysis with DPM1 and DPM2 yielded an Adjusted R^2^ 0.61 and 0.39 (Figure 2).

### Bacterial diseases

Prevalence of Whooping cough, Chlamydia, Diphtheria correlated negatively with DPM except UTI which correlated positively (Table1). Combining these parameters yielded Adjusted R2 of 0.41 and 0.39 with DPM1 and DPM2 respectively (Figure 2).

### Respiratory diseases

Upper respiratory infection correlated negatively while Interstitial lung disease and pulmonary sarcoidosis correlated positively (Table1). Regression analysis with DPM1 and DPM2 yielded an Adjusted R^2^ 0.20 and 0.19 respectively (Figure 2). Hence no significant association of respiratory diseases with deaths could be seen in our studies.

### Lifestyle diseases

High percentage of BMI (Body mass Index) deaths, High fasting plasma glucose, High systolic blood pressure correlated positively while a good percentage of child and maternal nutrition correlated negatively with DPM (Table1). Overall an adjusted R^2^ was achieve of 0.24 and 0.28 with DPM1 and DPM2 respectively (Figure 2). Hence we could not consider these values significant.

### Cancer

All cancers highly correlated with the DPM (Table1) and combining all cancer parameters for regression analysis with DPM1 and DPM2 yielded adjusted R^2^ of 0.45 and 0.64 respectively (Figure 2).

### Sanitation

Improved sanitation parameters across states such as percentage of Drinking water within premises, bathroom availability correlated positively with DPM1 with Closed drainage showing a very high rho= 0.67 and 0.71 for DPM1 and DPM2 respectively (Table1) while parameters lacking in sanitation such as percentage of bathing in enclosure without roof, no drainage and unsafe water sanitation and handwash deaths showed negative correlation (Table1). Adjusted R^2^ value for combined sanitation parameters was 0.62 and 0.61 with DPM1 and DPM2 respectively (Figure 2).

### Multivariate Linear Regression

Overall combining broader variables such as demography, sanitation, autoimmune diseases and cancer gave as adjusted R square value of 0.71 with DPM1 and 0.85 with DPM2 (Figure 2).

States with higher percentage of older population and with better sanitation parameters along with higher prevalence of autoimmune disorders along with cancer were more likely to face higher mortality.

## Discussion

A clear pattern of Covid-19 mortality has emerged globally, with wealthier nations experiencing higher mortality rates compared to lower middle and middle-income countries[1]. Our analysis of different Indian states also indicates a similar trend (Figure 1. (*a*)). It is well-established that economic prosperity has a significant impact on factors such as life expectancy, literacy, improved sanitations in a country. Our findings indicate that Indian states with higher GDP and better demographic parameters, such as higher literacy and life expectancy, have reported more Covid-related deaths (Figure 2).

The observation that states with a higher percentage of older population are positively correlated with DPM1 and DPM2 is in confirmation with other studies as the elderly are known to be more vulnerable to Covid-19[2,3]. Additionally, higher literacy rates in these states may contribute to more employment opportunities and mobility, which can increase exposure to the virus[22]. Furthermore, deaths are positively correlated with the level of urbanization in a state due to the higher likelihood of viral transmission in densely populated areas. We conclude that GSDP per capita does not directly contribute to Covid-19 deaths, as it may be a confounding variable for other developmental factors. Excluding GSDP per capita, the remaining developmental factors show a higher regression score with DPM1 and DPM2 (Figure 2), with very little contribution from GSDP per capita itself.

In our previous analysis[5], we had observed and is well-known that economic affluence in nations brings better sanitation parameters, such as safe drinking water, handwashing practices, closed toilets, and drainage systems. However, despite these improvements, these nations experienced a higher burden of Covid-19 mortality per million individuals. In our present analysis, we have observed a similar trend in Indian states, where parameters contributing to better sanitation, such as safe drinking water, toilets, and closed drainage within households, positively correlating with Covid-19 deaths.

One of our most perplexing observations was that of a strong correlation between deaths per million and closed drainage and/or availability of indoor toilets. Intuitively, availability of closed or open toilets should not have any effect on the spread of the virus, or mortality arising thereof, although fragments of RNA of the SARS-CoV-2 virus have been traced in drainage systems around the world[23,24]. One possible explanation for this observation is that Covid-19 being airborne can spread throughout an entire household through aerosols, including through toilets, even while patients are in isolation[10,11]. Studies suggest that flushing toilets can help suspend the virus in larger droplets, increasing air transmission and the infectivity of the virus[12,13]. We also note that states with open sanitation systems, without bathrooms, drainage, or roofs, tend to have lower Covid-19 death rates (Table 2). This may be due to a higher chance of the virus dissipating in the air and lower transmission to individuals.

**Table 1.** Shows the spearman correlation coefficient(rho),S-statistic(S) and p value(p-value) of explanatory variables with Covid-19 deaths/million in Indian states as on 17-09-2020 and 07-05-2021 respectively; N=28.

|  | Deaths per million as on 17-09-2020 |  |  | Deaths per million as on 07-05-2021 |  |  |
| --- | --- | --- | --- | --- | --- | --- |
| | $\rho$ | S | p-value | $\rho$ | S | p-value |
| **Explanatory Variables** |  |  |  |  |  |  |
| **Demography** |  |  |  |  |  |  |
| GSDP per capita(2018-2019) | 0.54 | 1696 | 0.004 | 0.66 | 1248 | 0.0002 |
| Age over 60(%) | 0.44 | 2060.47 | 0.02034 | 0.46 | 1979.4 | 0.01418 |
| Literacy(%) | 0.41 | 2138 | 0.02906 | 0.59 | 1500 | 0.00119 |
| Urbanisation(%) | 0.71 | 1074.65 | 0.00003 | 0.57 | 1567.71 | 0.00151 |
| Good Governance Index(2019) | 0.46 | 1963.77 | 0.01319 | 0.47 | 1922.76 | 0.01087 |
| Population below poverty(%) | -0.4 | 5113.2 | 0.03527 | -0.53 | 5599.27 | 0.00354 |
| Slum household(%) | 0.44 | 2053.56 | 0.01974 | 0.11 | 3263.89 | 0.5887 |
| **Sanitation** |  |  |  |  |  |  |
| Drinking water within premises(%) | 0.39 | 2236 | 0.04216 | 0.47 | 1938 | 0.01249 |
| Bathroom available(%) | 0.47 | 1926.53 | 0.01107 | 0.69 | 1126.31 | 0.00005 |
| Bathing in enclosure without roof(%) | -0.13 | 4128 | 0.50899 | -0.4 | 5122 | 0.03498 |
| Closed drainage(%) | 0.67 | 1212 | 0.00015 | 0.71 | 1070 | 0.00004 |
| No drainage(%) | -0.49 | 5444 | 0.00885 | -0.63 | 5950 | 0.00046 |
| Unsafe water sanitation and handwash deaths(%) | -0.38 | 5028 | 0.04941 | -0.62 | 5934 | 0.00051 |
| **Vaccination** |  |  |  |  |  |  |
| BCG(%) | 0.41 | 2160.59 | 0.03082 | 0.65 | 1283.35 | 0.00019 |
| POLIO(%) | 0.29 | 2584 | 0.13041 | 0.52 | 1756 | 0.00517 |
| DPT(%) | 0.35 | 2392.83 | 0.07205 | 0.58 | 1529.71 | 0.00118 |
| Measles(%) | 0.47 | 1926.76 | 0.01108 | 0.69 | 1122.65 | 0.00004 |
| Hepatitis(%) | 0.22 | 2868 | 0.2704 | 0.47 | 1942 | 0.01271 |
| Varicella and herpes zoster(%) | 0.36 | 2356 | 0.06427 | 0.59 | 1484 | 0.00108 |
| Acute hepatitis B(%) | -0.49 | 5460 | 0.00819 | -0.49 | 5440 | 0.00902 |
| Acute hepatitis E(%) | -0.30 | 4746 | 0.12241 | -0.50 | 5474 | 0.00765 |
| Vit A(%) | 0.51 | 1778 | 0.00579 | 0.51 | 1794 | 0.00627 |
| **Tropical diseases** |  |  |  |  |  |  |
| Trichuriasis(%) | -0.4 | 5114 | 0.03516 | -0.43 | 5232.33 | 0.02171 |
| Leprosy(%) | -0.44 | 5252 | 0.02085 | -0.59 | 5812 | 0.00116 |
| Hookworm(%) | -0.44 | 5274.47 | 0.01809 | -0.56 | 5703.77 | 0.0019 |
| Cystic echinococcus(%) | 0.19 | 2962.49 | 0.33479 | 0.45 | 2014.94 | 0.01666 |
| Ascaris(%) | -0.32 | 4826 | 0.0964 | -0.47 | 5382 | 0.01182 |
| **Autoimmune diseases** |  |  |  |  |  |  |
| Gout(%) | 0.4 | 2178 | 0.03393 | 0.56 | 1600 | 0.00219 |
| Psoriasis(%) | -0.5 | 5476 | 0.00758 | -0.63 | 5956 | 0.00044 |
| Diabetes mellitus type 2(%) | 0.4 | 2192 | 0.03579 | 0.53 | 1704 | 0.00393 |
| Asthma(%) | -0.51 | 5530 | 0.00579 | -0.64 | 5998 | 0.00032 |
| IBD(%) | 0.45 | 2002 | 0.01658 | 0.64 | 1316 | 0.00034 |
| **Viral diseases** |  |  |  |  |  |  |
| Measles(%) | -0.57 | 5733.28 | 0.00158 | -0.73 | 6309.36 | 0.00001 |
| Rabies(%) | -0.48 | 5414.24 | 0.00944 | -0.45 | 5306.23 | 0.0157 |
| Meningitis(%) | -0.46 | 5322 | 0.01546 | -0.59 | 5818 | 0.00112 |
| Acute hep(%) | -0.6 | 5848 | 0.00092 | -0.63 | 5968 | 0.0004 |
| Dengue(%) | 0.6 | 1470 | 0.00098 | 0.4 | 2208 | 0.03801 |
| **Bacterial diseases** |  |  |  |  |  |  |
| Whooping cough(%) | -0.51 | 5500 | 0.00673 | -0.65 | 6032 | 0.00025 |
| Diphtheria(%) | -0.44 | 5252 | 0.02085 | -0.64 | 5996 | 0.00033 |
| UTI(%) | 0.41 | 2174 | 0.03342 | 0.59 | 1504 | 0.00122 |
| Chlamydia(%) | -0.6 | 5858 | 0.00086 | -0.6 | 5860 | 0.00085 |
| **Respiratory diseases** |  |  |  |  |  |  |
| Upper respiratory infections(%) | -0.5 | 5482 | 0.00736 | -0.54 | 5640 | 0.00322 |
| Interstitial lung disease and pulmonary sarcoidosis(%) | 0.42 | 2108 | 0.0258 | 0.35 | 2378 | 0.06917 |
| **Lifestyle diseases** |  |  |  |  |  |  |
| Child maternal nutrition(%) | -0.54 | 5638 | 0.00326 | -0.7 | 6226 | 0.00005 |
| High bmi deaths(%) | 0.65 | 1278 | 0.00025 | 0.73 | 994 | 0.00002 |
| High fasting plasma<br>glucose(%) | 0.69 | 1144 | 0.00008 | 0.72 | 1016 | 0.00002 |
| High systolic blood<br>pressure(%) | 0.53 | 1704 | 0.00393 | 0.55 | 1636 | 0.00269 |
| **Cancer** |  |  |  |  |  |  |
| Breast cancer(%) | 0.46 | 1966 | 0.01416 | 0.44 | 2028 | 0.01854 |
| Brain and central nervous<br>system cancer(%) | 0.5 | 1834 | 0.00765 | 0.59 | 1512 | 0.00129 |
| Prostate cancer(%) | 0.35 | 2390 | 0.07196 | 0.61 | 1436 | 0.00078 |
| Kidney cancer(%) | 0.5 | 1816.5 | 0.00638 | 0.73 | 984.27 | 0.00001 |
| Hodgkin lymphoma(%) | 0.56 | 1610.72 | 0.00198 | 0.56 | 1591.72 | 0.00176 |
| Non Hodgkin<br>lymphoma(%) | 0.49 | 1874.54 | 0.00858 | 0.43 | 2067.74 | 0.02099 |

One key characteristic of developed states owing to better hygiene conditions is their lower incidences of communicable diseases such as helminths, parasitic and viral infections[16]. We have grouped them under tropical, viral and bacterial diseases. We find negative correlation of tropical diseases to deaths individually. However, combining them showed very low regression score with DPM1 and DPM2, this could be attributed again to the possibility that most states in India specially the urbanized area report very low rates of these parasitic diseases. The incidence of viral diseases and bacterial diseases individually correlated negatively with deaths with overall higher regression scores with DPM1 and DPM2. Studies suggest a protective humoral and cell mediated immunity being imparted to individuals with previous infections with viruses, bacteria and parasites that may last lifetime[25]. It has been suggested that exposure to infection helps dendritic cells to induce T cell regulation which work as a part of anti-inflammatory network and suppresses the cytokines[26,27]. Since majority of the Covid deaths and severity results from respiratory failure from acute respiratory distress syndrome (ARDS)[28] resulting from elevated inflammatory cytokines, it may be postulated that previous infection helps in keeping a check over a heightened immune response as compared to encountering lower infections. On the contrary, having autoimmune diseases may aggravate an already elevated inflammatory response in the body. In this aspect we see a positive correlation with gouty arthritis, diabetes and inflammatory bowel diseases. It is difficult to conclude if the immune reaction would be heightened or reduced as several patients may be under medication of immuno suppressives. As Asthma and Psoriasis are diseases that need constant medication it becomes apparent that they may not cause a very high reaction to antigens of Covid-19.

The respiratory diseases and lifestyle diseases show low scores of regression with DPM1 and DPM2 even though individually upper respiratory infections, BMI (Body mass Index) deaths, High fasting plasma glucose, high systolic blood pressure correlated positively with Covid deaths. On the other hand, different kinds of Cancers correlated positively with deaths as well as give a high regression score with DPM1 and DPM 2. Studies have shown cancer to be a high risk comorbidity factor conducted for individuals in several populations[8]. Cancer prevalence is also known to be higher with lifestyle associated with living in urban population[29,30].

It should be noted that while certain risk factors may be associated with higher or lower rates of Covid-related deaths, these associations do not necessarily indicate causality. Furthermore, population-level studies may not always reflect individual-level differences, which can vary due to genetic factors. Nevertheless, some correlations can be explained through existing domain knowledge and global trends observed in Covid patients, such as the impact of age, cancer, and lifestyle-related diseases on Covid severity. The prevalence of these lifestyle related diseases such as obesity, high blood pressure and Cancer are higher in developed countries as well as Indian States showing a high correlation with DPM1 and DPM2 (Table1). In other cases, we adopt a hygiene hypothesis-driven approach, recognizing the potential role of communicable and non-communicable diseases and sanitation in Covid outcomes. This can be very well seen with higher correlation of DPM1 and DPM2 with sanitation parameters, bacterial viral infections and the autoimmune disorders (Table1). We suggest exploring the possibility of microbiome therapy as a means of preventing future pandemics. Our studies can also aid in identifying states that require urgent government interventions to control Covid-19 mortality.

## Contributors

SCM conceived the study idea. BC collected the data, applied statistical tools and wrote the manuscript. Both the authors discussed the result interpretations and suggested correction to manuscript.

## Data sharing statement

The data used for analysis is publicly available. We have also enclosed our dataset used for analysis.

## Data Availability

All data produced in the present work are contained in the manuscript

## Acknowledgements

We thank Rajeeva Karandikar for several useful discussions. BC gratefully acknowledges financial support by the National Centre for Cell Science, Pune. Authors declare no competing financial interests. SCM acknowledges Anand Deshpande for philanthropic funding for Distinguished Professorship at the Savitribai Phule Pune University.

